# Changing of therapeutic trends between the 1st and 2nd wave did not reduce COVID-19 related mortality of renal transplant recipients: a national registry study

**DOI:** 10.1101/2021.12.15.21267794

**Authors:** Bastien Berger, Marc Hazzan, Nassim Kamar, Hélène Francois, Marie Matignon, Clarisse Greze, Philippe Gatault, Luc Frimat, Pierre F Westeel, Valentin Goutaudier, Renaud Snanoudj, Charlotte Colosio, Antoine Sicard, Dominique Bertrand, Christiane Mousson, Jamal Bamoulid, Antoine Thierry, Dany Anglicheau, Lionel Couzi, Jonathan M Chemouny, Agnes Duveau, Valerie Moal, Yannick Le Meur, Gilles Blancho, Jérôme Tourret, Paolo Malvezzi, Christophe Mariat, Jean-Philippe Rerolle, Nicolas Bouvier, the French SOT COVID Registry, Sophie Caillard, Olivier Thaunat

## Abstract

SARS-CoV-2 pandemic evolved in two consecutive waves over 2020 (for France: 1st wave from March 1 to July 31; and 2nd wave from August 1 to December 31). Improvements in the management of COVID-19 led to a reduction of mortality rates in hospitalized patients during the second wave. Whether this progress also benefited to kidney transplant recipients (KTR), a population particularly vulnerable to severe COVID-19, remained unclear.

In France, 957 KTR were hospitalized for COVID-19 in 2020 and their data were prospectively collected in the French SOT COVID registry. The presentation, management, and outcomes of the 359 KTR diagnosed during the 1st wave were compared to those of the 598 of the 2nd wave. Baseline comorbidities were largely similar between KTR of the 2 waves. Maintenance immunosuppression was reduced in most patients but withdrawal of antimetabolite (73.7% vs 58.4%, p<0.001) or CNI (32.1% vs 16.6%, p<0.001) was less frequent during the 2nd wave. Hydroxychloroquine and azithromycin that were commonly used during the 1st wave (21.7% and 30.9%, respectively) were almost abandoned during the 2nd. In contrast, the use of high dose corticosteroids doubled (19.5% vs. 41.6%, p<0.001). Despite these changing trends in COVID-19 management, 60-day mortality was not statistically different between the 2 waves (25.3% vs. 23.9%; Log Rank, p=0.48).

We conclude that changing of therapeutic trends during 2020 did not reduce COVID-19 related mortality in KTR. Our data indirectly support the importance of vaccination and monoclonal neutralizing anti-SARS-CoV-2 antibodies to protect KTR from severe COVID-19.

## Introduction

After the initial outbreak in China in late 2019, coronavirus disease (COVID-19) spread globally^1^. As on October 14, 2021, the pandemic had affected more than 238 million people causing more than 4.8 million deaths worldwide^2^.

Like in the rest of the world^3,4^, the viral pandemic evolved during 2020 in two consecutive waves in France. The first hit France in spring, only three months after SARS-CoV-2 discovery^5^, in a context of limited knowledge about COVID-19, absence of proven specific treatment, and shortage of essential equipment such as face masks and diagnostic tests^6,7^. The government imposed a national lockdown from March 17, 2020 to May 10, 2020, which successfully reduced the spread of the virus and led to the resolution of this first wave^8^. However, in the absence of available vaccine, SARS-CoV-2 resurged following the ease of social distancing rules during the summer. As a result, a second pandemic wave started in fall 2020. However, in contrast with the first wave, enhanced testing capacities allowed diagnosis of asymptomatic cases during this second wave. Additionally, intensivists had better experience of the stereotypical course of severe COVID-19, including the prolonged mechanical ventilation and Intensive Care Unit (ICU) stay^9^, the increased risk of thrombotic events^10^, and the high rates of acute kidney injury^11^. More importantly, RECOVERY trial^12^ had been published, providing evidence that Dexamethasone reduces mortality in hospitalized patients requiring oxygen therapy by one fifth. These changes in medical care resulted in a 10% reduction of mortality rates in French hospitalized patients during the second wave compared to the first one^13,14^.

Whether kidney transplant recipients (KTR), a particularly vulnerable population to COVID-19^15–17^, also benefited from the progresses made along 2020 in COVID-19 management remained unclear. Aiming at addressing this question, we retrospectively analyzed the prospectively collected data of the French Solid Organ Transplant (SOT) COVID registry and compare the course, management and outcomes of COVID-19 diagnosed in the first versus second waves in 957 hospitalized French KTR.

## Methods

### Data Collection

KTR hospitalized for COVID-19 in France between March 1 and December 31 2020 were identified by the interrogation of the French Solid Organ Transplant (SOT) COVID registry.

This prospective registry was approved by the Institutional Review Board of Strasbourg University (approval number 02.26) and registered at clinicaltrials.gov (NCT04360707). Of note, all patients were informed about their inclusion in the registry but the need for informed consent was waived.

### Study Design and Patients

Inclusion criteria were age > 18 years at the diagnosis of COVID-19 and presence of a functioning kidney graft. The diagnostic criteria for COVID-19 was based on: (i) a positive RT-PCR for SARS-CoV-2 in nasopharyngeal swab or (ii) the presence of typical respiratory symptoms accompanied by evocative pulmonary lesions on low-dose chest CT when RT-PCR yielded negative results. Cases were considered to have occurred during the 1^st^ wave if they were diagnosed between March 1 to July 31, 2020 and during 2^nd^ wave if they were diagnosed between August 1 to December 31, 2020. We used the time cutoff of December 31, 2020 for the end of the second wave to have an equal length of time compared to the first wave and to avoid the effect of the vaccination to increase baseline comparability. Cardiovascular disease included heart failure, coronary vascular disease and dysrhythmia. Respiratory disease included chronic respiratory failure, asthma and chronic obstructive pulmonary disease.

### Statistical Analysis

Categorial variables are reported as counts and percentages. Continuous variables are presented as medians and interquartile ranges. Differences between groups were assessed with the χ^2^ test or 2-sided Fisher’s exact test for categorical variables and with Student’s t-test or Wilcoxon’s rank-sum test for continuous variables. Survival curves were represented using the Kaplan-Meier method and compared with the log-rank test. The primary outcome is 60-day mortality. Secondary outcomes are: admission to the ICU, 60-day mortality in ICU, initiation of renal replacement therapy (RRT), use of mechanical ventilation, use of vasopressor support, occurrence of bacterial pulmonary superinfection, or thrombo-embolic event. The multiple imputations method^18^ was used to handle missing data on relevant covariates. Five imputed data sets were generated and analyses were performed on each of them. Then, the results were combined using the Rubin rules^19^ to obtain average values. To assess risk factors for mortality, Cox proportional hazard univariable and multivariable models were built. All the variables with a univariable threshold p<0.1 were selected as covariates for the initial multivariable model. The covariates in the final multivariable model were selected using a backward conditional procedure with a threshold p<0.05. Results are expressed as hazard ratios (HRs) with their 95% confidence intervals. All analyses were conducted in the R environment (R Foundation for Statistical Computing, Vienne, Austria) version 4.1.2^20^ using the “survival” and “mice” packages. All tests were 2-sided, and p<0.05 was considered statistically significant.

## Results

### Baseline patient characteristics

Shortage in diagnosis assays during the first pandemic wave resulted in the fact that only symptomatic patients were tested to confirm clinically or radiologically suspected COVID-19^21,22^. As the result of enhanced availability of these assays along the year 2020, asymptomatic COVID-19 were identified during the second wave^21^. Furthermore, from January 2021 onward anti-SARS-CoV-2 vaccines became available, reducing the risk of severe COVID-19 and contributing to the resolution of the second pandemic wave. Since the criteria for hospitalization of KTR with symptomatic COVID-19 little evolved over time and given the fact that our aim was to compare the two pandemic waves, the present study focused on the 957 cases [n=359 (37.5%) from the first and n=598 (62.5%) from the second wave] of COVID-19 diagnosed in KTR that require hospitalization and occurred before January 1^st^ 2021.

The characteristics of enrolled patients, which were prospectively collected in the French SOT COVID registry, are presented in **Table 1**. Briefly, a little less than 10% of the cohort received a graft from a living donor. The median recipient age was 63.0 [52.0-70.0] years and males represented 68.1% of the cohort. The majority of patients (537/864, 62.1%) were overweight and the median BMI of the cohort was 26.0 [23.0-29.4] kg/m^2^. The most common comorbidity was hypertension (798/918, 86.9%), followed by diabetes (371/914, 40.6%) and cardiovascular disease (352/908, 38.8%). The median baseline estimated glomerular filtration rate (eGFR) was 41.0 [30.0-54.0] mL/min/1.73m^2^. Regarding therapeutic immunosuppression, the vast majority of patients received an induction therapy, either with anti-interleukin-2 (385/931, 41.4%) or with antithymocyte globulin (508/931, 54.6%). At diagnosis of COVID-19, maintenance regimen of most patients consisted in a combination of calcineurin inhibitor (807/957, 84%, either tacrolimus 65.3% or cyclosporine 19%), an antimetabolite (722/957, 75.4% on mycophenolic acid) and corticosteroids (726/957, 75.9%). Only 4.0% of the cohort were on belatacept.

**TABLE 1.**
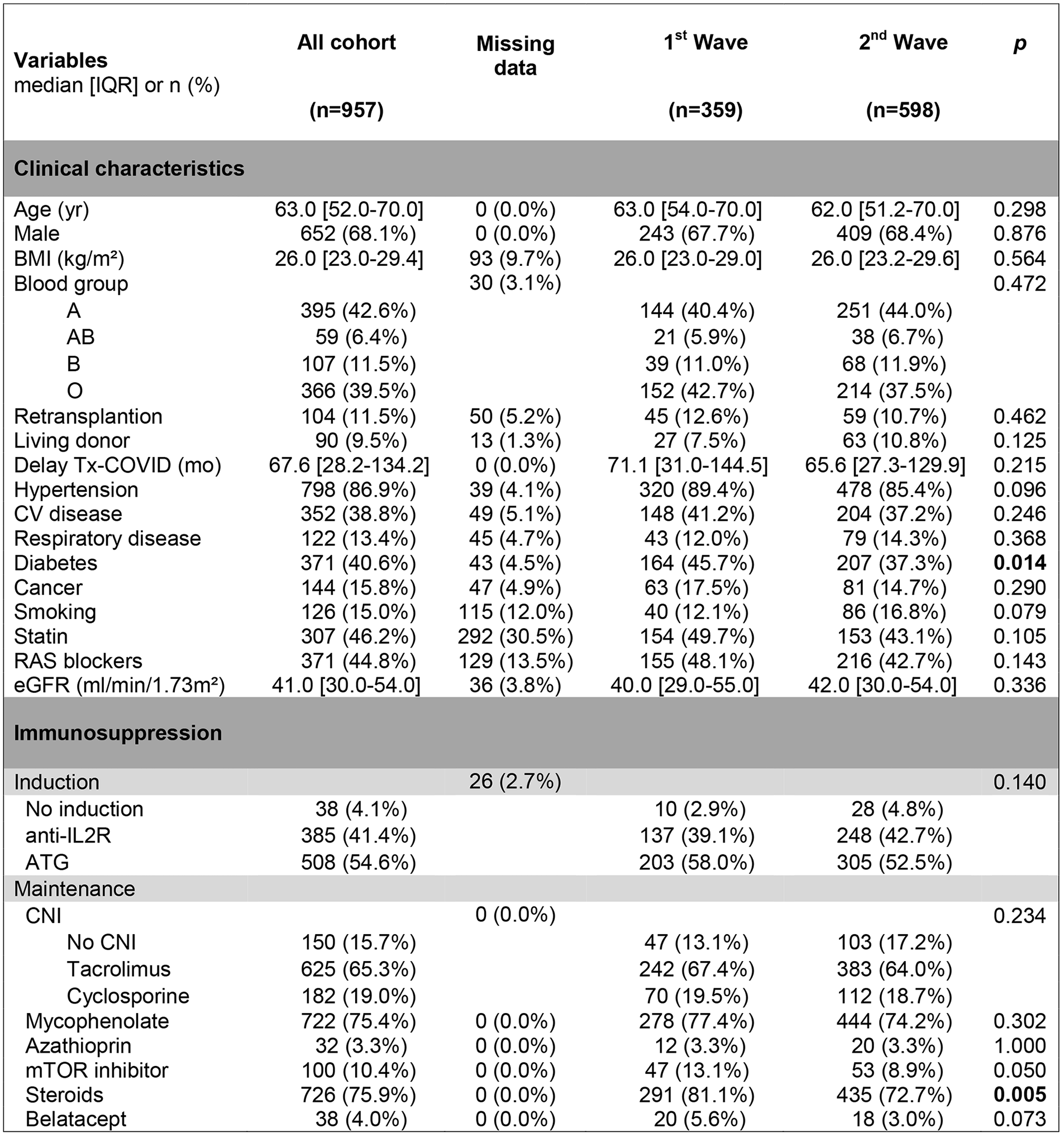
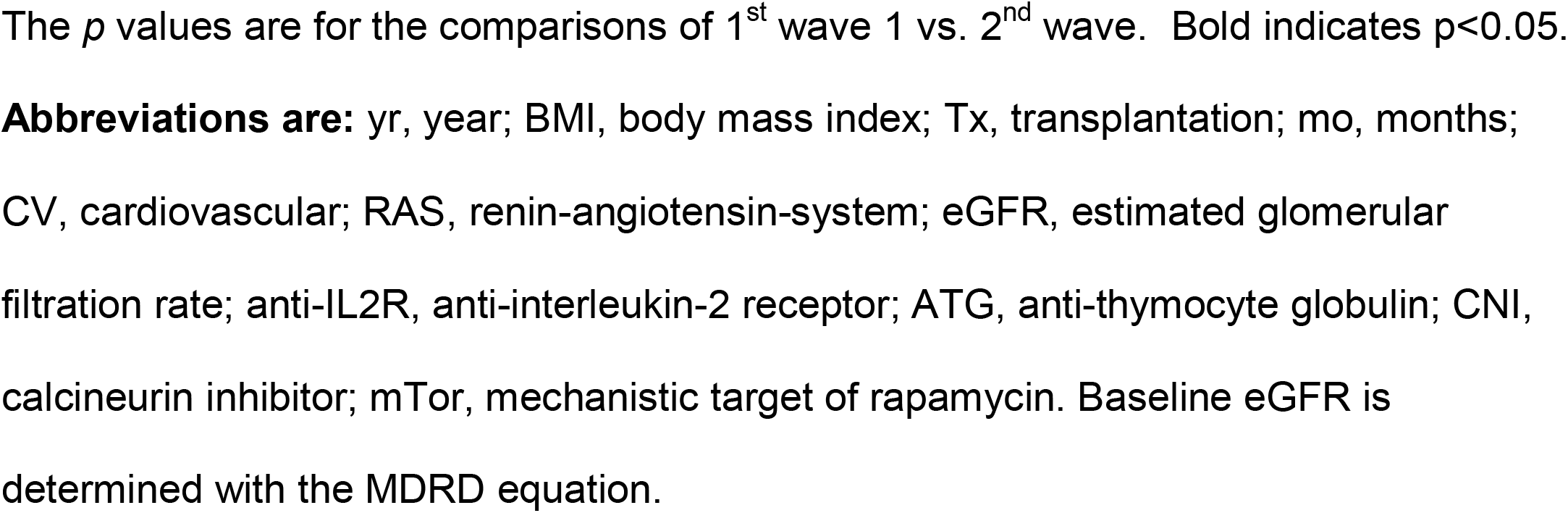
Baseline characteristics of kidney transplant patients at admission for COVID-19.

| Variables<br>median [IQR] or n (%) | All cohort<br>(n=957) | Missing<br>data | 1<sup>st</sup> Wave<br>(n=359) | 2<sup>nd</sup> Wave<br>(n=598) | p |
| --- | --- | --- | --- | --- | --- |
| **Clinical characteristics** |  |  |  |  |  |
| Age (yr) | 63.0 [52.0-70.0] | 0 (0.0%) | 63.0 [54.0-70.0] | 62.0 [51.2-70.0] | 0.298 |
| Male | 652 (68.1%) | 0 (0.0%) | 243 (67.7%) | 409 (68.4%) | 0.876 |
| BMI (kg/m<sup>2</sup>) | 26.0 [23.0-29.4] | 93 (9.7%) | 26.0 [23.0-29.0] | 26.0 [23.2-29.6] | 0.564 |
| Blood group |  | 30 (3.1%) |  |  | 0.472 |
| A | 395 (42.6%) |  | 144 (40.4%) | 251 (44.0%) |  |
| AB | 59 (6.4%) |  | 21 (5.9%) | 38 (6.7%) |  |
| B | 107 (11.5%) |  | 39 (11.0%) | 68 (11.9%) |  |
| O | 366 (39.5%) |  | 152 (42.7%) | 214 (37.5%) |  |
| Retransplantation | 104 (11.5%) | 50 (5.2%) | 45 (12.6%) | 59 (10.7%) | 0.462 |
| Living donor | 90 (9.5%) | 13 (1.3%) | 27 (7.5%) | 63 (10.8%) | 0.125 |
| Delay Tx-COVID (mo) | 67.6 [28.2-134.2] | 0 (0.0%) | 71.1 [31.0-144.5] | 65.6 [27.3-129.9] | 0.215 |
| Hypertension | 798 (86.9%) | 39 (4.1%) | 320 (89.4%) | 478 (85.4%) | 0.096 |
| CV disease | 352 (38.8%) | 49 (5.1%) | 148 (41.2%) | 204 (37.2%) | 0.246 |
| Respiratory disease | 122 (13.4%) | 45 (4.7%) | 43 (12.0%) | 79 (14.3%) | 0.368 |
| Diabetes | 371 (40.6%) | 43 (4.5%) | 164 (45.7%) | 207 (37.3%) | **0.014** |
| Cancer | 144 (15.8%) | 47 (4.9%) | 63 (17.5%) | 81 (14.7%) | 0.290 |
| Smoking | 126 (15.0%) | 115 (12.0%) | 40 (12.1%) | 86 (16.8%) | 0.079 |
| Statin | 307 (46.2%) | 292 (30.5%) | 154 (49.7%) | 153 (43.1%) | 0.105 |
| RAS blockers | 371 (44.8%) | 129 (13.5%) | 155 (48.1%) | 216 (42.7%) | 0.143 |
| eGFR (ml/min/1.73m<sup>2</sup>) | 41.0 [30.0-54.0] | 36 (3.8%) | 40.0 [29.0-55.0] | 42.0 [30.0-54.0] | 0.336 |
| **Immunosuppression** |  |  |  |  |  |
| Induction |  | 26 (2.7%) |  |  | 0.140 |
| No induction | 38 (4.1%) |  | 10 (2.9%) | 28 (4.8%) |  |
| anti-IL2R | 385 (41.4%) |  | 137 (39.1%) | 248 (42.7%) |  |
| ATG | 508 (54.6%) |  | 203 (58.0%) | 305 (52.5%) |  |
| **Maintenance** |  |  |  |  |  |
| CNI |  | 0 (0.0%) |  |  | 0.234 |
| No CNI | 150 (15.7%) |  | 47 (13.1%) | 103 (17.2%) |  |
| Tacrolimus | 625 (65.3%) |  | 242 (67.4%) | 383 (64.0%) |  |
| Cyclosporine | 182 (19.0%) |  | 70 (19.5%) | 112 (18.7%) |  |
| Mycophenolate | 722 (75.4%) | 0 (0.0%) | 278 (77.4%) | 444 (74.2%) | 0.302 |
| Azathioprin | 32 (3.3%) | 0 (0.0%) | 12 (3.3%) | 20 (3.3%) | 1.000 |
| mTOR inhibitor | 100 (10.4%) | 0 (0.0%) | 47 (13.1%) | 53 (8.9%) | 0.050 |
| Steroids | 726 (75.9%) | 0 (0.0%) | 291 (81.1%) | 435 (72.7%) | **0.005** |
| Belatacept | 38 (4.0%) | 0 (0.0%) | 20 (5.6%) | 18 (3.0%) | 0.073 |

The *p* values are for the comparisons of 1<sup>st</sup> wave 1 vs. 2<sup>nd</sup> wave. Bold indicates $p < 0.05$ .
**Abbreviations are:** yr, year; BMI, body mass index; Tx, transplantation; mo, months; CV, cardiovascular; RAS, renin-angiotensin-system; eGFR, estimated glomerular filtration rate; anti-IL2R, anti-interleukin-2 receptor; ATG, anti-thymocyte globulin; CNI, calcineurin inhibitor; mTor, mechanistic target of rapamycin. Baseline eGFR is determined with the MDRD equation.

The patients of the two pandemic waves were largely similar except for diabetes, the prevalence of which was slightly lower in patients of the second wave (37.3% versus 45.7% respectively; p=0.014). Difference in immunosuppression regimen were also minor with only less patients on corticosteroids (72.7% vs 81.1%, p=0.005) and mTOR inhibitors (8.9% vs 13.1%, p=0.050) in the second pandemic wave.

### Clinical and biological presentation of COVID-19 at admission

Almost all diagnoses of COVID-19 (919/957, 96%) were confirmed by reverse transcriptase-polymerase chain reaction. SARS-CoV-2 infection occurred after a median of 67.6 [28.2-134.2] months after kidney transplantation. Of note, despite the fact that KT activity in France was interrupted during the first wave but maintained during the 2^nd^ there was no difference in the median delay from transplantation to COVID-19 diagnosis between the two pandemic waves (71.1 [31.0-144.5] vs 65.6 [27.3-129.9] months, p=0.215).

Considering the whole cohort (**Figure 1A**), the most frequent symptom on admission was fever (585/957, 67.2%), followed by cough (494/957, 56.8%), dyspnea (466/957, 52.3%) and diarrhea (317/957, 36.2%). Median levels of C-reactive protein and procalcitonin were 67 [28-121] mg/L and 0.22 [0.12-0.70] ng/mL respectively. At admission, most (580/653, 89%) patients had low lymphocyte count (median lymphocyte count of the cohort 0.65.10^9^ [0.40-1.00]/L) and median creatininemia was 174 [129-256] µmol/L.

**FIGURE 1.**
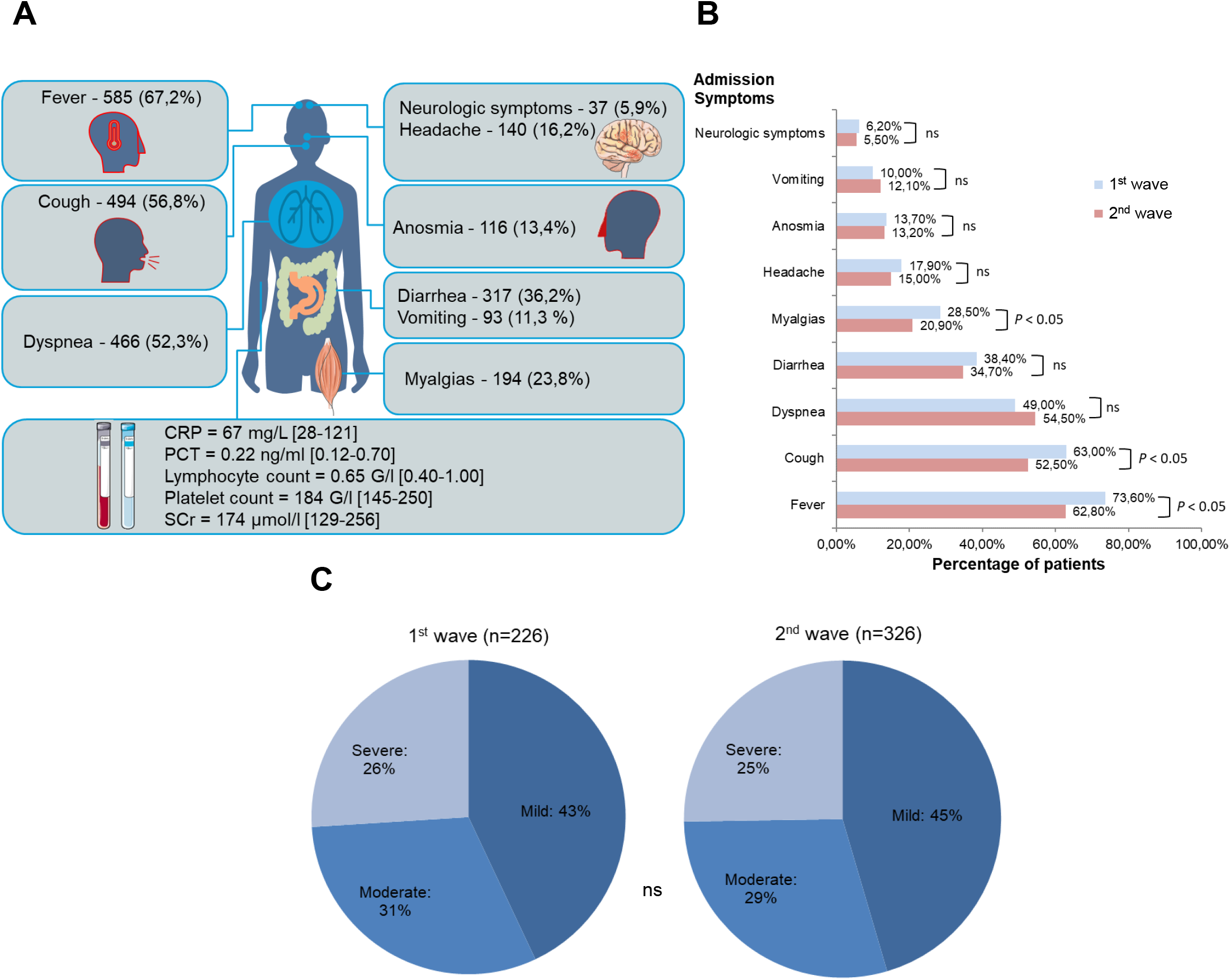
Clinical and biological presentation of COVID-19 at admission. **A** Summary of the main clinical and biological characteristics of the entire cohort (n=957 KTR), Median [IQR] or n (%), at hospital admission for COVID-19. **B** Comparison of characteristics at hospital admission for COVID-19 of patients from the 1^st^ vs 2^nd^ pandemic wave. **C** Comparison of chest CT scan severity between the 1^st^ vs 2^nd^ pandemic wave. **Abbreviations are:** CRP, C-reactive protein; PCT, procalcitonin; SCr, serum creatinine. *χ*^2^ test; p>0.05, ns.

KTR from the second wave differed from those of the first in that they less frequently exhibited fever, cough and myalgias, which could indicate earlier diagnosis during the second wave (**Figure 1B**). This hypothesis is coherent with the increased availability of diagnosis assays during the second half of 2020. However, no significant differences in CRP and PCT levels, nor in lymphocyte count could be observed between the 2 pandemic waves (data not shown). Furthermore, chest CT scan severity at presentation was also similar between the two waves with ∼45%, 30% and 25% of KTR presented with mild, moderate and severe degree of involvement respectively (**Figure 1C;** p=0.921).

### Management of immunosuppression and anti-viral therapies

The exact impact of maintenance immunosuppression during COVID-19 is unclear^23^. On one hand these drugs could be protective against the overproduction of proinflammatory cytokines during critical COVID-19^24,25^, on the other solid organ transplant recipients have been found to have delayed SARS-CoV-2 clearance^26,27^.

Maintenance immunosuppression was tapered in KTR hospitalized for symptomatic COVID-19, particularly antimetabolites and mTOR inhibitors, which were discontinued in the majority of patients of both pandemic waves (**Figure 2A**). However, if modifications of maintenance immunosuppression did not differ in nature between the two waves, they were made in a smaller proportion of patients during the second wave, particularly regarding withdrawal of CNI (32.1% vs 16.6%, p<0.001) and of antimetabolites (73.7% vs 58.4%, p<0.001; **Figure 2A**), which is in line with a previous report from the USA^28^. Contrasting with the global stability of immunosuppression management, anti-SARS-CoV-2 therapies differed in many respects between the two waves (**Figure 2B**). KTR with COVID-19 from the second wave received less frequently probabilistic antibiotics compared to those of the first wave (75.8% vs 49.2%, p<0.001). Hydroxychloroquine and azithromycin, which were commonly used during the first wave were almost completely abandoned during the second (21.7% vs 1.7% and 30.9% vs 5.0%, p<0.001, respectively). Tocilizumab use declined between the first and second waves (7.5% vs 2.2%, p<0.001). Conversely, the use of high dose corticosteroids doubled (19.5% vs. 41.6%, p<0.001).

**FIGURE 2.**
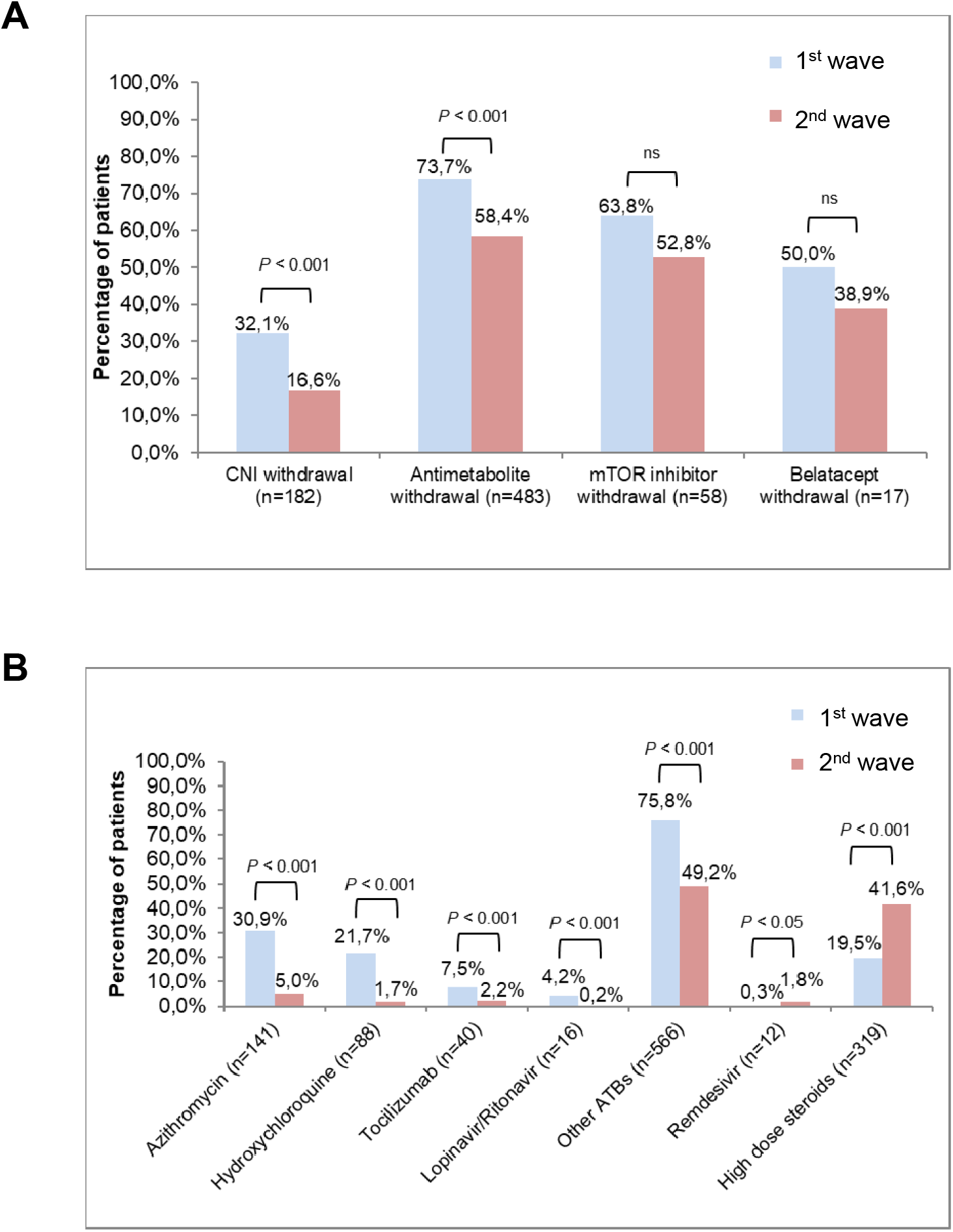
Changing of therapeutic trends between the 1st and 2^nd^ pandemic COVID-19 wave. Comparison of the management of immunosuppression (**A**) and the use of COVID-19 specific treatments (**B**) between the 1^st^ (blue) vs 2^nd^ (red) pandemic wave. **Abbreviations are:** CNI, calcineurin inhibitor; mTor, mechanistic target of rapamycin; ATB, antibiotics. *χ*^2^ test; p>0.05, ns.

### Risk factors associated with death due to COVID-19 in KTR

Univariate analysis conducted on the whole cohort identified: age, hypertension, preexisting cardiovascular disease, history of cancer, diabetes, dyspnea at admission, CRP > 60 mg/L at admission, baseline eGFR as significantly associated with mortality (data not shown). In contrast, diarrhea, anosmia and headaches were associated with reduced risk of death.

In multivariable analysis, only age>50 years, history of cancer, dyspnea or CRP > 60 mg/L at admission, and baseline eGFR<30ml/min/m^2^ remained independently associated with a higher risk of death in KTR hospitalized for COVID-19 (**Figure 3**), while anosmia at admission was associated with a better prognosis (**Figure 3**).

**FIGURE 3.**
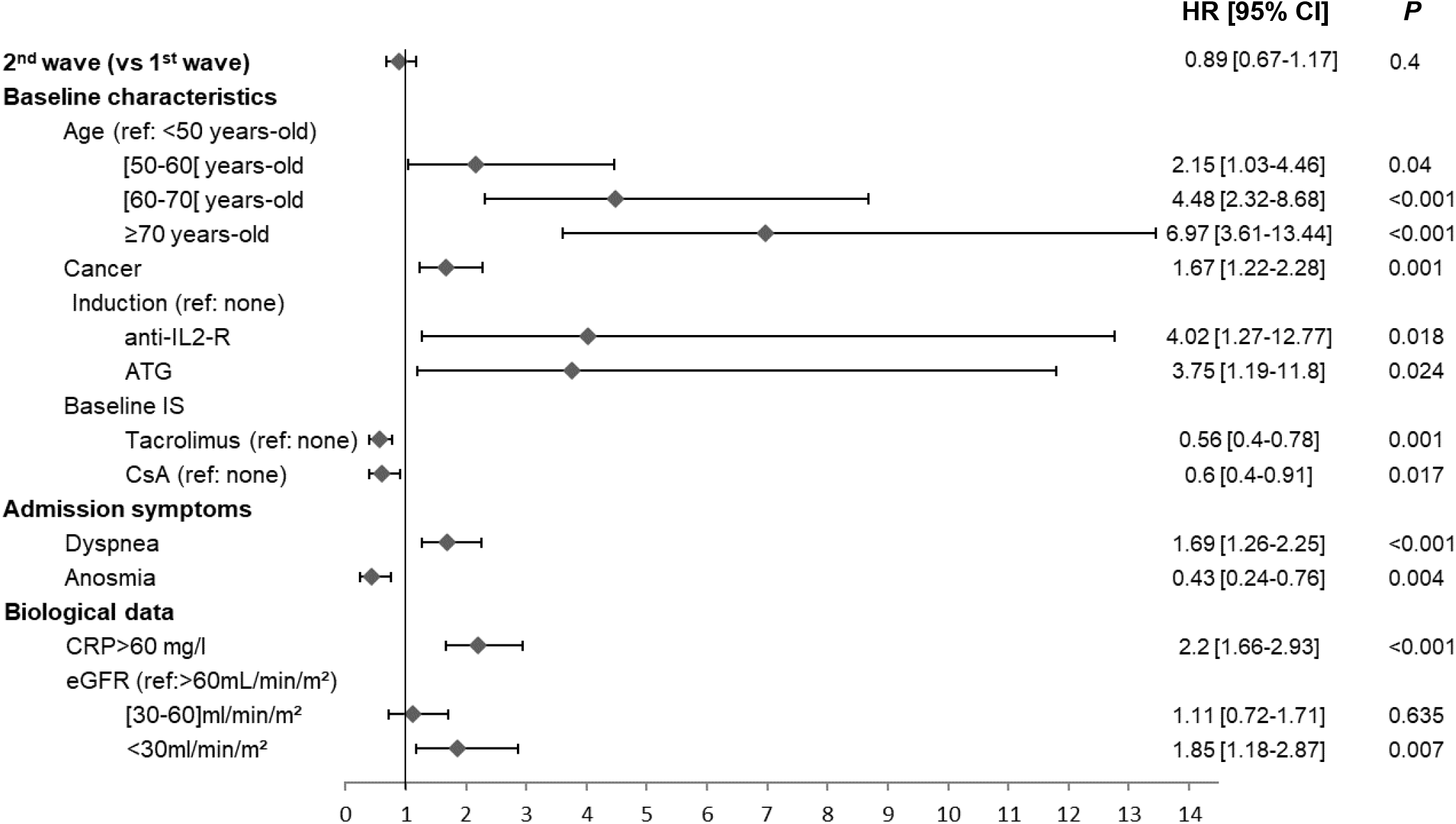
Variables associated with the risk of death due to COVID-19 in KTR. This forest plot shows the variable independently associated with the risk of death in multivariate analysis for the 957 KTR diagnosed with COVID-19 during the 1^st^ or the 2^nd^ pandemic wave.

### Comparison of first vs second wave outcomes

The incidence of thromboembolic events (9.5% vs 6.4%, p=0.135) and bacterial superinfection (27.0% vs 30.7%, p=0.304) was similar between the 2 pandemic waves. A non-significant trend for lesser use of RRT (15.9% vs 12.9%, p=0.230), mechanical ventilation (26.5% vs 22.1%, p=0.152) and vasopressor support (20.5% vs 15.9%, p=0.304) was observed during the 2^nd^ wave but mortality at 60 days from admission (24.5%) was in the range of what previously reported^29,30^, with no significant difference between the first and second wave (**Figure 4A**; Log rank test, p=0.48).

**FIGURE 4.**
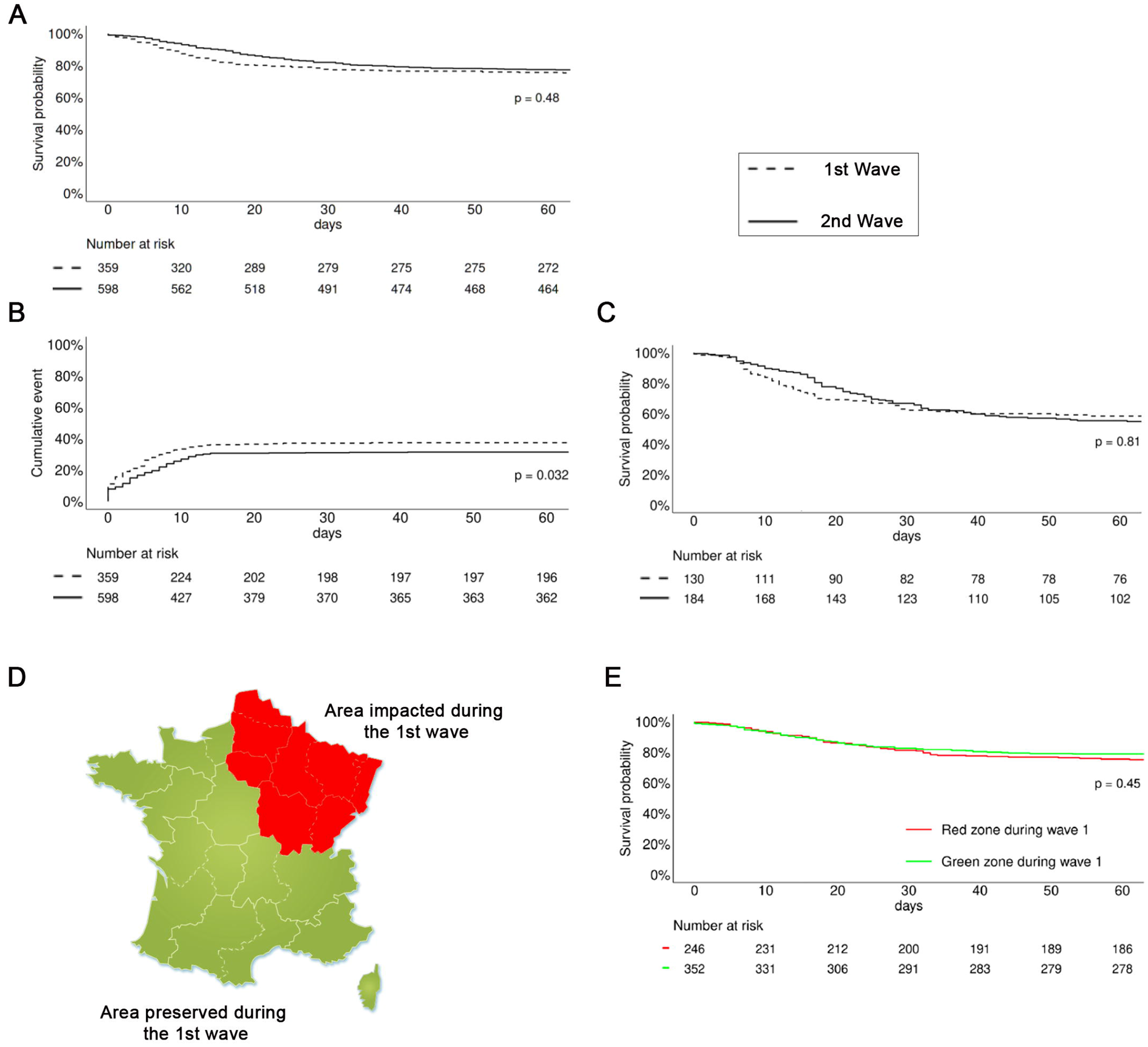
Comparison of COVID-19 outcomes between the 1^st^ and 2^nd^ wave. **A** In-hospital survival of KTR diagnosed with COVID-19 during the 1^st^ and 2^nd^ wave. **B** Cumulative incidence of Intensive Care Unit (ICU) admission of KTR diagnosed with COVID-19 during the 1^st^ and 2^nd^ wave. **C** Survival of KTR diagnosed with COVID-19 transferred in ICU. **D** Map of the geographic distribution of the cases of COVID-19 in France during the 1^st^ wave. Area in which the incidence of COVID-19 was the highest are in red. **E** In-hospital survival of KTR diagnosed with COVID-19 during the 2^nd^ wave according to their geographic location (in the red or green area defined in panel D). Comparison were made using the Log Rank test.

A slight difference in dynamic between the two waves could however be observed on Kaplan Meier curves (**Figure 4A**), with shorter delay between admission and death due to COVID-19 in KTR of the first wave (death at 14 days: 16.7% vs 10.1%, p=0.002). This difference is to be interpreted together with a faster and higher incidence of transfer in ICU for patients of the first wave (**Figure 4B**), without difference on the mortality for patients transferred in ICU (**Figure 4C**). Altogether, these findings could indicate that patients of the first wave were diagnosed (and therefore hospitalized) later in the course of COVID-19, a hypothesis in line with the difference in clinical presentation between the 2 waves reported above (**Figure 1B**).

In contrast with the second wave that impacted the entire French territory, the first pandemic wave had a heterogeneous geographic distribution^31^ that could have introduce a “learning-curve” bias. Physicians from the geographic area impacted by the first wave could have accumulated knowledge and skills useful to better manage patients from the second wave. To test this hypothesis, we compared the survival of KTR hospitalized for COVID-19 during the second wave in geographic area impacted (in red on the map **Figure 4D**) vs preserved (in green on the map **Figure 4D**) during the first pandemic wave. The similarity in survival for patients of the second wave hospitalized in either of these two areas strongly argue against the theory of the learning curve bias (**Figure 4E**).

## Discussion

Kidney transplant recipients (KTR), who are characterized by a highly comorbid profile and receive therapeutic immunosuppression to prevent graft rejection, were very early identified as particularly vulnerable to COVID-19^15–17^. An excess of mortality, integrally explained by COVID-19, was indeed reported in this population during the first wave of the pandemic in France^31^ and several large multicenter cohorts KTR estimated short-term intra-hospital mortality around 20-32%^29,32,33^. Among the risk factors identified in previous publications for death due to COVID-19 in KTR are age, eGFR and presence of comorbidities, including cardiovascular disease, diabetes, and/or obesity^31,32,34,35^. Additionally, dyspnea and elevations of biochemical markers of inflammation at diagnosis of COVID-19 were also associated with less favorable survival figures^36–38^. Our study largely confirms these data. In addition, it provides original additional information regarding the stability of the risk of death due to COVID-19 in KTR, despite the impressive accumulation of knowledge regarding the disease, which translated in better outcomes in the general population^13,14,39^. Indeed, despite a more homogeneous COVID-19 management with wider prescription of Dexamethasone and important decrease in the use of treatments deemed inefficient such as azithromycin^40^, hydroxychloroquine^40,41^, lopinavir/ritonavir^42^, survival of hospitalized KTR during the second wave remained similar to that observed during the first wave. Our findings concur with a meta-analysis including 5559 KTR with COVID-19 that reported a mean mortality rate of 23% (similar to what we observed) without significant difference between “early” (studies submitted before July 2020) and “late” (studies submitted from July 2020 onwards) phases of the pandemic^43^. These conclusions conflict with a recent study showing a better prognosis in “late” (from June 20 to December 31, 2020) compared to “early” 2020 (from March 1 to June 19, 2020) among 973 solid organ transplant recipients (SOTR) hospitalized in USA for COVID-19^28^. In their report, crude mortality by 28 days indeed declined from 19.6% in early period to 13.7% in late period and after adjusting for differences in baseline comorbidities between both periods, the odds of death remained lower in the late period (aOR 0.67, 95% CI 0.46 – 0.98, *p = 0*.*04*). Instead of the changing trends in management of COVID-19 patients, we believe that the observations made by Heldman et al, could be explained by the numerous differences in the baseline comorbid profiles of SOTR between the early and late period (SOTR in late period presented with less hypertension, diabetes, heart failure, coronary artery disease, chronic lung disease) and/or by the short follow-up period of the study. Indeed, when we assessed 14-days mortality in our own cohort, we also found a significant difference between the first and second wave (16.7% vs. 10.1%, p-value = 0.002, respectively) that progressively disappeared by the end of the 60-days follow-up period. Whether this effect is attributable to earlier diagnosis of COVID-19 in KTR during the second wave is possible and supported by some clues discussed above but remains to be formerly demonstrated.

Among the strengths of our study are the relative high number of patients enrolled and the prospective collection of data. Our study has however also some limitations. First, we compared two periods (first and second wave) but did not take into account COVID-19 ICU occupancy rates, a factor thought to impact on mortality rates^13^. Second, our study was not designed to capture the impact of vaccines, which only became available early 2021.

Accumulating evidence suggests however that KTR have an impaired response to the “standard” 2-dose of mRNA vaccine^44–47^, which leaves them at high risk of severe COVID-19^46,48^. Despite intensified scheme of vaccination (with third and even a fourth vaccine dose now recommended in weak responders), up to 20% of KTR will not develop sufficient protection against COVID-19^49–51^. In this regard, the development of monoclonal neutralizing anti-SARS-CoV-2 Spike Protein Antibodies represent an interesting therapeutic option. The latter are already available in high-risk patients diagnosed with mild to moderate COVID-19^52^ (post-exposition therapy) and first reports about their use for prophylaxis (pre-exposition therapy) are promising^53^.

In conclusion, changing of therapeutic trends during 2020 did not reduce COVID-19 related mortality in KTR. Our data thus indirectly stress the importance of therapeutic progresses made during 2021, including vaccination and monoclonal neutralizing anti-SARS-CoV-2 spike protein antibodies, to protect this vulnerable population from death due to COVID-19.

## Data Availability

All data produced in the present study are available upon reasonable request to the authors

## Acknowledgements

OT is supported by the Etablissement Français du Sang and the Fondation pour la Recherche Médicale (PME20180639518).

## Appendix

The French SOT COVID Registry Collaborators are as follows: Sophie Caillard, Bruno Moulin, Service de Néphrologie et Transplantation, Hôpitaux Universitaires de Strasbourg, Strasbourg; Samira Fafi-Kremer, Laboratoire de Virologie, Hôpitaux Universitaires de Strasbourg, Strasbourg; Marc Hazzan, Service de Néphrologie, Hôpital Huriez, Lille; Dany Anglicheau, Service de Néphrologie et Transplantation Adultes, AP-HP, Hôpital Necker, Paris; Alexandre Hertig, Jérôme Tourret, Benoit Barrou, Service de Néphrologie, AP-HP, Hôpital La Pitié Salpétrière, Paris; Emmanuel Morelon, Olivier Thaunat, Service de Néphrologie, Hôpital Edouard Herriot, Lyon; Lionel Couzi, Pierre Merville, Service de Néphrologie–Transplantation– Dialyse, Hôpital Pellegrin, Bordeaux; Valérie Moal, Tristan Legris, Service de Néphrologie et Transplantation, AP-HM, Hôpital de la Conception, Marseille; Pierre-François Westeel, Maïté Jaureguy, Service de Néphrologie, CHU Amiens Picardie, Amiens; Luc Frimat, Service de Néphrologie, CHRU Nancy, Vandoeuvre; Didier Ducloux, Jamal Bamoulid, Service de Néphrologie, Hôpital Jean-Minjoz, Besancon; Dominique Bertrand, Service de Néphrologie, CHU de Rouen, Rouen; Michel Tsimaratos, Florentine Garaix-Gilardo, Service de Pédiatrie Multidisciplinaire, Hôpital La Timone, Marseille; Jérôme Dumortier, Service d’Hépato-Gastroentérologie, Hôpital Edouard Herriot, Lyon; Sacha Mussot, Antoine Roux, Centre Chirurgical Marie Lannelongue, Le Plessis Robinson; Laurent Sebbag, Service d’Insuffisance Cardiaque, Hôpital Louis Pradel, Bron; Yannick Le Meur, Service de Néphrologie, Hôpital de la Cavale Blanche, Brest; Gilles Blancho, Christophe Masset, Service de Néphrologie–Transplantation, Hôtel Dieu, Nantes; Nassim Kamar, Service de Néphrologie et Transplantation, Hôpital Rangueil, Toulouse; Hélène Francois, Eric Rondeau, Service de Néphrologie, Dialyse et Transplantation, AP-HP, Hôpital Tenon, Paris; Nicolas Bouvier, Service de Néphrologie, Dialyse, Transplantation Rénale, CHU, Caen; Christiane Mousson, Service de Néphrologie, Dijon; Matthias Buchler, Philippe Gatault, Service de Néphrologie, Tours; Jean-François Augusto, Agnès Duveau, Service de Néphrologie, Dialyse, Transplantation, CHU Angers, Angers; Cécile Vigneau, Marie-Christine Morin, Jonathan Chemouny, Leonard Golbin, Service de Néphrologie, CHU de Rennes, Rennes; Philippe Grimbert, Marie Matignon, Antoine Durrbach, Service de Néphrologie, Hôpital Henri-Mondor, Creteil; Clarisse Greze, Service de Néphrologie, AP-HP, Hôpital Bichat Claude Bernard, Paris; Renaud Snanoudj, Service de Néphrologie, Hôpital Foch, Service de Néphrologie et Transplantation Hôpital du Kremlin Bicêtre, Le Kremlin Bicetre; Charlotte Colosio, Betoul Schvartz, Service de Néphrologie, Hôpital Maison Blanche, Reims; Paolo Malvezzi, Service de Néphrologie, Hémodialyse, Transplantation Rénale, Hôpital La Tronche, Grenoble; Christophe Mariat, Service de Néphrologie, CHU de Saint Etienne, Saint Etienne; Antoine Thierry, Service de Néphrologie, Hémodialyse et Transplantation Rénale, Hôpital Jean Bernard, Poitiers; Moglie Le Quintrec, Service de Néphrologie-Transplantation-Dialyse, CHU Lapeyronie, Montpellier; Antoine Sicard, Service de Néphrologie, Hôpital Pasteur, Nice; Jean Philippe Rerolle, Service de Néphrologie, CHU Dupuytren, Limoges; Anne-Élisabeth Heng, Cyril Garrouste, Service de Néphrologie, CHU Gabriel Montpied, Clermont-Ferrand; Henri Vacher Coponat, Service de Néphrologie, CHU de La Réunion, Saint Denis; Éric Epailly, Service de Cardiologie, Hôpitaux Universitaires de Strasbourg, Strasbourg; Olivier Brugiere, Service d’Hépatologie, Hôpital Foch, Suresnes; Sébastien Dharancy, Service d’Hépatologie, Hôpital Huriez, Lille; Éphrem Salame, Service de Chirurgie Hépatique, Hôpital Universitaire de Tours, Tours; Faouzi Saliba, Service d’Hépatologie, Centre hépato-biliaire Paul Brousse, Villejuif, France.

